# Improving MASLD Identification in Patients with T2D: A Computable Phenotype Framework Integrating Structured Data and Clinical Notes

**DOI:** 10.64898/2026.09.26.26364078

**Authors:** Yuanhan Chen, Anu Sharma, Qiang Yang, Yuyao Chen, Rui Yin

## Abstract

Metabolic dysfunction-associated steatotic liver disease (MASLD) is highly prevalent among individuals with type 2 diabetes (T2D), yet accurate identification from electronic health records (EHRs) remains challenging because clinically relevant information is fragmented across structured data and unstructured clinical documentation. We developed a rule-based computable phenotype (CP) framework that integrates structured EHRs with clinical notes for scalable MASLD identification in patients with T2D. The framework comprises five complementary CP algorithms derived from validated fibrosis indices and liver injury biomarkers, together with a lightweight natural language processing module that identifies MASLD-related evidence using keyword matching, contextual filtering, and negation detection. The proposed framework was systematically evaluated across multiple threshold settings, with additional validation through age- and sex-stratified analyses and manual chart review. The findings demonstrate that integrating structured and unstructured EHR data provides an accurate and scalable framework for real-world MASLD phenotyping and offers a practical strategy for developing CPs for other chronic diseases.

## Introduction

Metabolic dysfunction–associated steatotic liver disease (MASLD), formerly known as non-alcoholic fatty liver disease (NAFLD), is the most common chronic liver disease worldwide, affecting nearly one-quarter of the adult population (1–4), with an estimated global incidence rate of approximately 4.6 new cases per 100,000 person-years (5). MASLD is closely associated with obesity, insulin resistance, and type 2 diabetes mellitus (T2D), reflecting its role as the hepatic manifestation of systemic metabolic dysfunction (6). Individuals with T2D are disproportionately affected, with approximately 60–70% developing MASLD and a substantially increased risk of progression to advanced fibrosis, cirrhosis, hepatocellular carcinoma, and liver-related mortality compared with individuals without diabetes (7–9). Moreover, the coexistence of MASLD and T2D is associated with poorer glycemic control, increased cardiovascular risk, and higher all-cause mortality (10,11), highlighting the close interplay between metabolic dysfunction and chronic liver disease. Early and accurate identification of MASLD in patients with T2D is therefore essential for timely risk stratification, targeted intervention, and prevention of progressive liver disease.

Despite its clinical importance, accurately identifying MASLD in real-world healthcare settings remains challenging. Definitive diagnosis typically relies on liver biopsy or liver-specific imaging, both of which are not routinely available for large-scale population surveillance. Consequently, computable phenotypes (CPs) have emerged as an effective approach for scalable disease identification by translating clinical knowledge into reproducible, rule-based algorithms that can be implemented across diverse electronic health record (EHR) systems (12,13). While structured EHR data, such as laboratory measurements and diagnosis codes, provide standardized and readily accessible information (14), they frequently fail to capture the complete clinical picture. Unstructured clinical narratives and imaging reports often contain valuable diagnostic evidence unavailable in coded data. Integrating structured and unstructured EHR data is therefore critical for improving the completeness, sensitivity, and robustness of EHR-based phenotyping (15,16).

Recent efforts have developed CP frameworks to identify MASLD using EHRs. Geyer et al. demonstrated that rule-based phenotyping using laboratory fibrosis indices and metabolic parameters can identify substantial numbers of under-coded MASLD cases in large clinical populations (17). Similarly, Liu et al. proposed and validated a structured-data CP algorithm that achieved high accuracy and specificity for MASLD (18). However, these approaches primarily depended on structured EHR variables and did not systematically leverage complementary evidence embedded in clinical narratives, potentially overlooking patients whose diagnostic information is documented only in free-text notes. These limitations highlight the need for multimodal CP frameworks that integrate structured and unstructured EHR data to improve case ascertainment while maintaining interpretability and scalability.

In this study, we developed and evaluated a multimodal CP framework for MASLD identification in patients with T2D by integrating structured EHRs and clinical text. The proposed framework combined five complementary rule-based phenotyping strategies derived from established fibrosis indices, liver enzyme measurement, and metabolic risk scores with contextual information extracted from clinical narratives. We systematically evaluated multiple threshold settings, quantified the contribution of note-derived evidence, and assessed performance across demographic subgroups. By combining complementary EHR modalities within an interpretable and reproducible framework, this study provides a scalable approach for real-world MASLD phenotyping and offers a generalizable strategy for developing CPs for other chronic diseases.

## Methodology

### Overall Study Design

This study developed and evaluated a rule-based CP framework for identifying MASLD in patients with T2D using both structured and unstructured EHR data. First, five MASLD CP rules were constructed based on established clinical guidelines and published phenotyping algorithms, incorporating laboratory biomarkers, liver enzyme measurements, fibrosis indices, and metabolic risk scores. Multiple threshold values were evaluated for each rule to identify clinically appropriate operating points in a real-world EHR setting. Second, unstructured clinical notes were involved as complementary information and evidence to enhance MASLD identification. Finally, the proposed framework was evaluated using standard diagnostic performance metrics and subgroup analyses stratified by age and sex. To further assess validity and generalizability, an independent set of 240 patients with T2D were reviewed through manual chart annotation, and the resulting labels were compared with CP-derived classifications to assess agreement and identify discrepancies.

### Data Source and Cohort Construction

We utilized clinical data from the University of Florida (UF) Health Integrated Data Repository (IDR) that involves both structured EHR data, e.g., demographics, diagnostic codes and laboratory measurements, and unstructured clinical notes. As illustrated in **Figure 1**, a total of 109,393 patients with T2D were initially identified from UF Health IDR. Candidate MASLD cases were subsequently identified using predefined ICD-9/10 diagnosis codes (ICD-9: 571.8, 571.9; ICD-10: K75.81, K76.0), yielding 17,681 patients with documented MASLD and 91,712 patients without documented MASLD.

**Figure 1.**
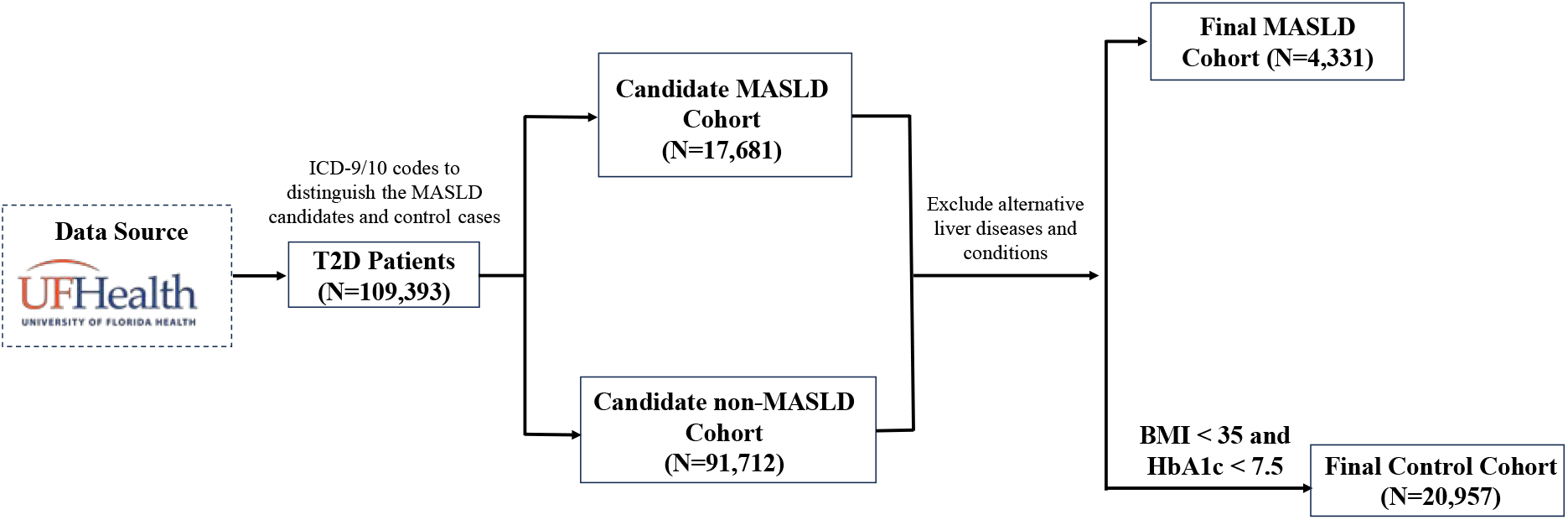
Cohort construction workflow. Patients with T2D were identified from the UF Health Integrated Data Repository and stratified into candidate MASLD and non-MASLD cohorts using ICD-9/10 diagnosis codes. Sequential application of exclusion criteria and additional metabolic eligibility criteria yielded the final study and control cohorts.

Following initial cohort identification, identical exclusion criteria were applied to both the candidate MASLD and non-MASLD cohorts to remove patients with alternative liver diseases or other conditions that could confound MASLD identification. Patients with alternative liver diseases or conditions that could confound MASLD identification were excluded using predefined ICD-9/10 diagnosis codes, including alcohol-related liver disease (K70), chronic viral hepatitis (B18 and B19), autoimmune hepatitis (K75.4), inflammatory liver diseases (K75.0-K75.3, K75.9), Wilson disease (E83.0), hemochromatosis (E83.11) and cystic fibrosis (E84). After applying these exclusion criteria, the candidate MASLD cohort was reduced from 17,681 to 4,331 patients, forming the final study cohort.

For the candidate non-MASLD cohort, additional eligibility criteria were applied to reduce the likelihood of including individuals with undiagnosed or subclinical MASLD. Specifically, patients with a body mass index (BMI) ≥30 kg/m^2^ or hemoglobin A1c (HbA1c) ≥7.5% were further excluded because obesity and poor glycemic control are well-established risk factors for MASLD and could indicate patients with unrecognized disease despite the absence of diagnostic codes. After these additional filtering steps, 20,957 unique patients remained and constituted the final control cohort.

This study complied with all relevant ethical regulations and was approved by the University of Florida Institutional Review Board (protocol no. IRB202300939). All patients’ clinical data were de-identified, and the institution waived the requirement for written informed consent. This study used no human biological samples and was conducted in accordance with the principles of the 2024 Declaration of Helsinki.

### Rule-Based Computable Phenotype Development

Five rule-based CP algorithms were constructed based on established laboratory biomarkers and validated clinical metrics relevant to MASLD. These rules capture complementary aspects of disease pathology, including hepatocellular injury, metabolic dysfunction, and hepatic fibrosis. The corresponding formulas and clinical thresholds were performed directly within the EHR-based phenotyping framework. Each rule was implemented as an independent CP algorithm and evaluated separately to characterize its diagnostic performance before integration with unstructured clinical notes.

#### (i) Rule 1: Fibrosis-4 (FIB-4) Index

The FIB-4 index (19) is a widely used non-invasive fibrosis score that estimates hepatic fibrosis using age, aspartate aminotransferase (AST), alanine aminotransferase (ALT), and platelet count (PLT). It is calculated as: 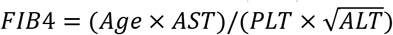. FIB-4 has been extensively validated across multiple international guidelines as a reliable non-invasive predictor of advanced hepatic fibrosis and a practical tool for risk stratification in patients with steatotic liver disease. It has also been applied in large-scale EHR studies for identifying patients at increased risk of MASLD-related fibrosis (17).

#### (ii) Rule 2: ALT Elevation

ALT is a commonly used biomarker of hepatocellular injury and has been widely incorporated into MASLD screening and phenotyping algorithms (20,21). Persistent ALT elevation has demonstrated utility for identifying patients with suspected MASLD in population-scale EHR studies.

#### (iii) Rule 3: AST Elevation

Similarly to ALT, AST is another established indicator of hepatocellular injury and fibrosis progression. Elevated AST levels have been associated with increasing disease severity in MASLD-related fibrosis and complement ALT in identifying hepatic injury (22).

#### (iv) Rule 4: Fibrosis-Related Index (FNI)

FNI is a composite fibrosis score integrating liver enzymes and metabolic biomarkers, calculated as *FNI* = 1/(1 + *e*^−*x*^), where *x* is the sum of clinical markers and constants formulated as *x* = −10.33 + 2.54 * *ln*(*AST*) + 3.86 * *ln*(*HbA*1*c*) − 1.66 * *ln*(*HDL*). By combining biochemical indicators of liver injury and metabolic dysfunction, FNI provides a noninvasive assessment of fibrosis risk (23).

#### (iv) Rule 5: NAFLD Score

The NAFLD Score is a validated noninvasive fibrosis assessment that integrates demographic, metabolic, and laboratory parameters, including age, body mass index (BMI), diabetes status, the AST/ALT ratio, platelet count, and albumin (24). It has been widely adopted for identifying patients at increased risk of advanced hepatic fibrosis. Although originally developed for NAFLD, the score remains applicable under the MASLD nomenclature as it relies on metabolic and hepatic biomarkers central to disease progression (25).

### Clinical Notes Processing and Integration

To enhance the identification of MASLD beyond structured EHR data, clinical notes were incorporated as an additional information source. A rule-based natural language processing (NLP) pipeline was applied to identify MASLD-related concepts from clinical notes (20,21). A curated terminology lexicon was constructed based on commonly used clinical expressions and disease nomenclature, including “fatty liver,” “steatosis,” “steatohepatitis,” “NAFLD,” “NASH,” “MAFLD,” “MASH,” and “non-alcoholic liver disease.” Keyword matching was performed at the encounter level, consistent with the EHR-based CP framework. To improve precision, a multi-stage validation process was implemented. First, keyword matches associated with negated statements were excluded using predefined negation patterns (e.g., “*no evidence of*”, “*denies*”, “*negative for*”, “*without*”, “*free of*”) occurring within the local textual context of each keyword (20,21). Second, contextual cue words such as “*impression*”, “*assessment*”, “*diagnosis*”, and “*consistent with*” were used to prioritize clinically meaningful diagnostic statements over incidental or descriptive mentions. Finally, keyword matches referring to family history or other non-patient contexts (e.g., “*history of*”, “*family history of*”) were excluded to avoid misclassification (20,21). Following contextual filtering, each encounter containing at least one valid MASLD-related keyword was assigned a positive note-derived indicator. During CP identification, the structured EHR-based rule and the note-derived indicator were integrated using a logical OR strategy, whereby an encounter was classified as MASLD if it satisfied either the structured EHR-based CP rule or the note-based criterion. This integration allowed clinically documented evidence contained in free-text narratives to complement structured EHR-based rules, improving case ascertainment while preserving the interpretability of the rule-based phenotyping framework.

### Validation and Evaluation

For each of the five rule-based CP algorithms, multiple threshold values were evaluated to characterize diagnostic performance. This threshold analysis enabled systematic comparison of sensitivity-specificity tradeoffs and identification of clinically appropriate thresholds for subsequent analyses. Diagnostic performance was quantified using sensitivity, specificity, positive predictive value (PPV), and F1-score. To assess the contribution of unstructured clinical information, each CP rule was evaluated under two configurations: (1) structured EHR data alone and (2) structured EHR data integrated with the clinical note-derived indicator. Performance differences between the two configurations were used to quantify the incremental value of incorporating free-text clinical documentation into the phenotyping framework. The proposed CP framework was further evaluated using an independent manually reviewed cohort, which served as the reference standard for MASLD status. In addition, subgroup analyses were performed by age and sex using the selected threshold for each rule to assess the robustness and consistency of the framework across demographic groups.

## Results

### Cohort summary and characteristics

Table 1 summarizes the baseline demographic and clinical characteristics of the study and control cohorts. The study cohort comprised 4,331 unique patients with both T2D and MASLD, while the control cohort included 20,957 participants with T2D only. Compared with the control cohort, individuals with MASLD were younger on average (55.31vs. 62.65 years) and had a higher proportion of females (64.9% vs. 53.3%). The racial and ethnic distributions were generally comparable between the two cohorts, with non-Hispanic White individuals representing the largest subgroup in both populations (69.1% vs. 65.3%). Patients with MASLD also exhibited a higher mean body mass index (35.62 vs. 28.06 kg/m^2^) but a slightly lower mean high-density lipoprotein (HDL) cholesterol level (44.59 vs. 49.10 mg/dL). These baseline characteristics demonstrate distinct demographic and metabolic differences between the MASLD and control cohorts.

**Table 1.** Descriptive statistics on the characteristics of the study and control cohorts.

|  |  |  |
| --- | --- | --- |
| **Characteristics** | **Study Cohort**<br>**Patients with both T2D**<br>**and MASLD (n=4,331)** | **Control Cohort**<br>**Patients with T2D only**<br>**(n=20,957)** |
| **Demographics** |  |  |
| **Age, mean (years)** | 55.31 | 62.65 |
| **Male, N (%)** | 1,519 (35.1%) | 9,783 (46.7%) |
| **Female, N (%)** | 2,812 (64.9%) | 11,174 (53.3%) |
| **Race/Ethnicity** |  |  |
| **Hispanics** | 416 (9.6%) | 1,557 (7.4%) |
| **Non-Hispanic White** | 2,994 (69.1%) | 13,675 (65.3%) |
| **Non-Hispanic Black** | 599 (13.8%) | 4,012 (19.1%) |
| **Other** | 315 (7.3%) | 1,515 (7.2%) |
| **Unknown** | 7 (0.2%) | 198 (0.9%) |
| **BMI, mean (kg/m<sup>2</sup>)** | 35.62 (8.63) | 28.06 (4.35) |
| **HDL Cholesterol, mean (mg/dL)** | 44.59 (13.63) | 49.10 (15.73) |
| **Characteristics** | **Study Cohort**<br>**Patients with both T2D**<br>**and MASLD (n=4,331)** | **Control Cohort**<br>**Patients with T2D only**<br>**(n=20,957)** |
| **HbA1c, mean (%)** | 6.87 (1.57) | 6.20 (0.67) |
| **AST, mean (IU/L)** | 47.46 (38.65) | 28.73 (23.43) |
| **ALT, mean (IU/L)** | 42.62 (41.69) | 25.74 (25.89) |

### Performance of MASLD Computable Phenotype Rules

The performance of the five MASLD CP rules was evaluated across multiple threshold settings using structured EHR data only (Table 2). For each rule, a representative threshold was selected to achieve relatively balanced diagnostic performance while prioritizing sensitivity for identifying potentially under-coded MASLD cases. Among all the CP rules, FIB-4 showed moderate performance across threshold settings. At a threshold of FIB-4 > 1.0, the rule achieved an accuracy of 0.539, sensitivity of 0.563, specificity of 0.512, and an F1-score of 0.561. Lower thresholds further increased sensitivity but were accompanied by substantial reductions in specificity. The liver enzyme-based rules exhibited more consistent performance. Using sex-specific thresholds (ALT: male >28 IU/L, female >18 IU/L), the ALT rule achieved an accuracy of 0.683, sensitivity of 0.705, specificity of 0.661, and an F1-score of 0.696. Under the same thresholds, the AST rule achieved an accuracy of 0.637, sensitivity of 0.665, specificity of 0.607, and an F1-score of 0.653. The FNI rule provided the highest sensitivity among the evaluated methods at more permissive thresholds. At FNI > 0.1, it achieved an accuracy of 0.685, sensitivity of 0.751, specificity of 0.619, and the highest F1-score (0.705). In contrast, the NAFLD Score exhibited greater variability across threshold settings than the other rules. Higher thresholds favored specificity at the expense of sensitivity, whereas a threshold of NAFLD Score > -0.5 produced a more balanced performance, with an accuracy of 0.584, sensitivity of 0.601, specificity of 0.566, and an F1-score of 0.601. Overall, the liver enzyme-based (ALT and AST) and FNI rules demonstrated stronger overall performance than FIB-4 and the NAFLD Score, providing balanced sensitivity and specificity for rule-based MASLD identification using structured EHR data.

**Table 2.**
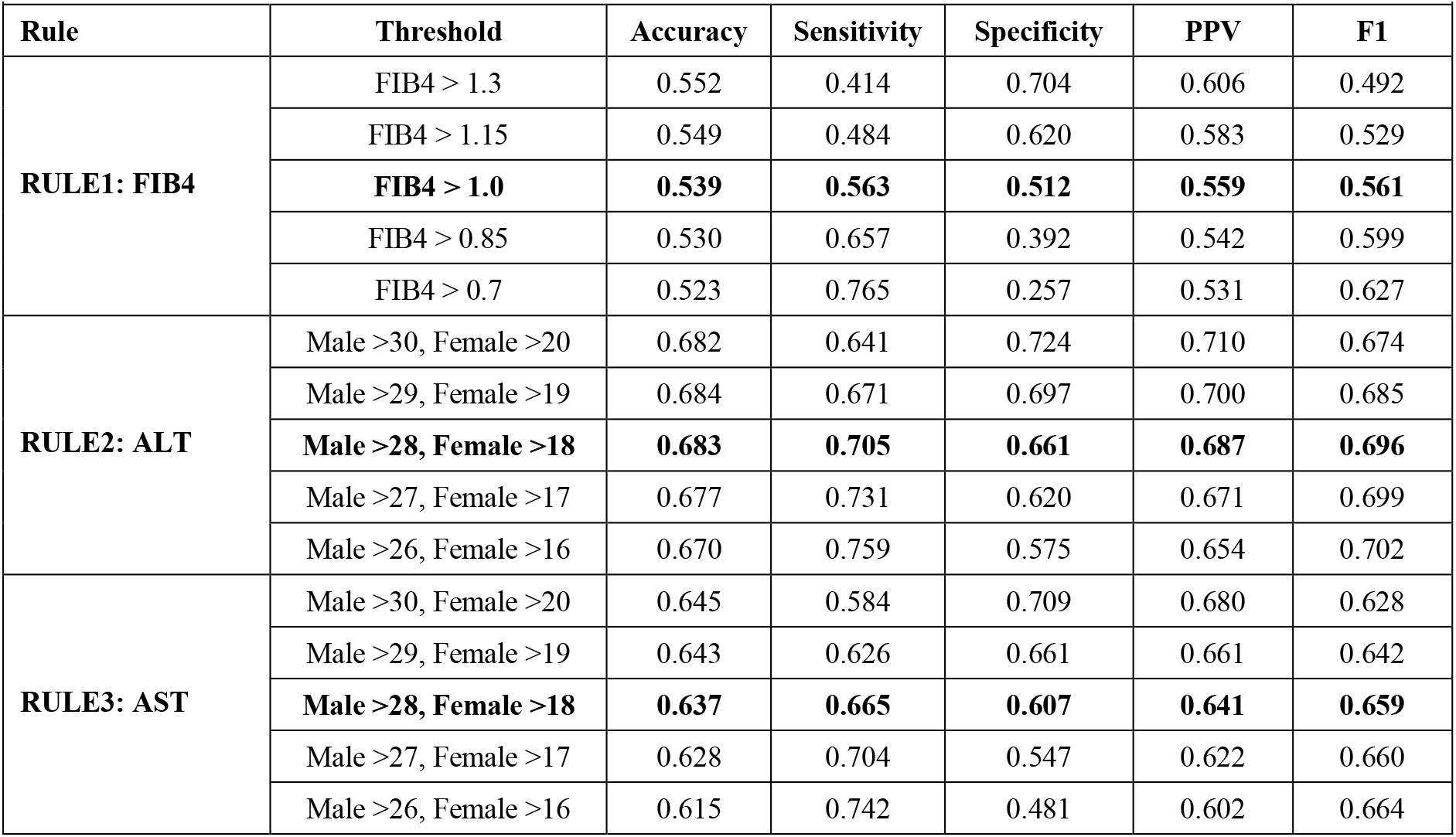
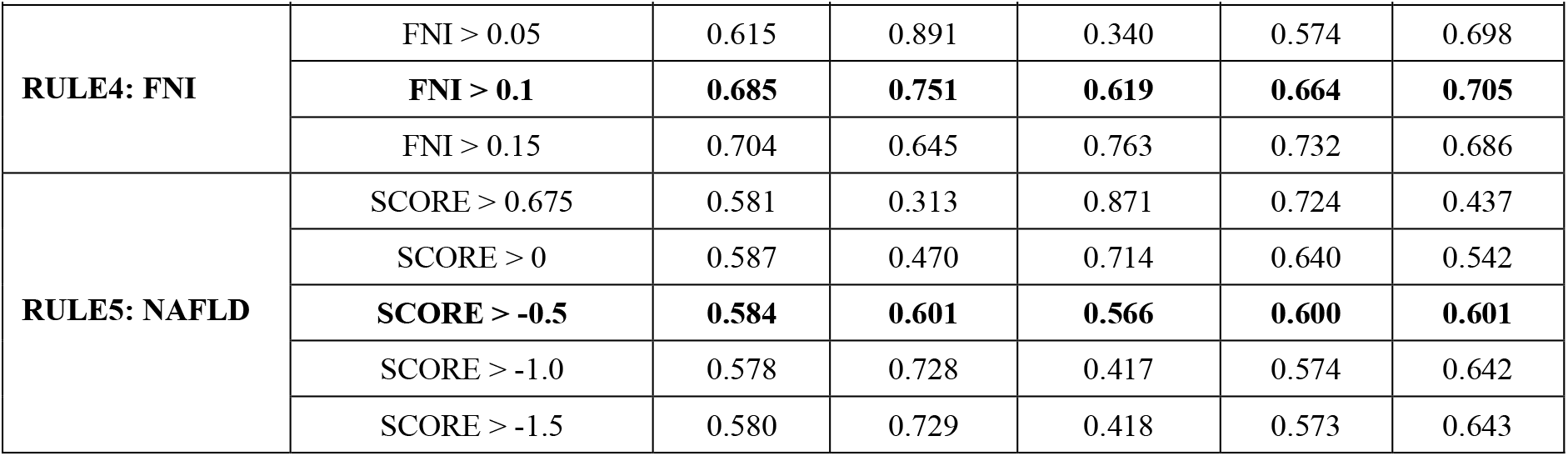
Performance of the five rule-based CPs for MASLD across multiple threshold settings using structured EHR data alone. Diagnostic performance is reported in terms of accuracy, sensitivity, specificity, positive predictive value (PPV), and F1-score. Bold rows indicate the threshold selected for subsequent analyses based on balanced diagnostic performance.

|  |  |  |  |  |  |  |
| --- | --- | --- | --- | --- | --- | --- |
| **Rule** | **Threshold** | **Accuracy** | **Sensitivity** | **Specificity** | **PPV** | **F1** |
| **RULE1: FIB4** | FIB4 > 1.3 | 0.552 | 0.414 | 0.704 | 0.606 | 0.492 |
|  | FIB4 > 1.15 | 0.549 | 0.484 | 0.620 | 0.583 | 0.529 |
|  | **FIB4 > 1.0** | **0.539** | **0.563** | **0.512** | **0.559** | **0.561** |
|  | FIB4 > 0.85 | 0.530 | 0.657 | 0.392 | 0.542 | 0.599 |
|  | FIB4 > 0.7 | 0.523 | 0.765 | 0.257 | 0.531 | 0.627 |
| **RULE2: ALT** | Male >30, Female >20 | 0.682 | 0.641 | 0.724 | 0.710 | 0.674 |
|  | Male >29, Female >19 | 0.684 | 0.671 | 0.697 | 0.700 | 0.685 |
|  | **Male >28, Female >18** | **0.683** | **0.705** | **0.661** | **0.687** | **0.696** |
|  | Male >27, Female >17 | 0.677 | 0.731 | 0.620 | 0.671 | 0.699 |
|  | Male >26, Female >16 | 0.670 | 0.759 | 0.575 | 0.654 | 0.702 |
| **RULE3: AST** | Male >30, Female >20 | 0.645 | 0.584 | 0.709 | 0.680 | 0.628 |
|  | Male >29, Female >19 | 0.643 | 0.626 | 0.661 | 0.661 | 0.642 |
|  | **Male >28, Female >18** | **0.637** | **0.665** | **0.607** | **0.641** | **0.659** |
|  | Male >27, Female >17 | 0.628 | 0.704 | 0.547 | 0.622 | 0.660 |
|  | Male >26, Female >16 | 0.615 | 0.742 | 0.481 | 0.602 | 0.664 |

| Rule | Threshold | Accuracy | Sensitivity | Specificity | PPV | F1 |
| --- | --- | --- | --- | --- | --- | --- |
| RULE4: FNI | FNI > 0.05 | 0.615 | 0.891 | 0.340 | 0.574 | 0.698 |
|  | **FNI > 0.1** | **0.685** | **0.751** | **0.619** | **0.664** | **0.705** |
|  | FNI > 0.15 | 0.704 | 0.645 | 0.763 | 0.732 | 0.686 |
| RULE5: NAFLD | SCORE > 0.675 | 0.581 | 0.313 | 0.871 | 0.724 | 0.437 |
|  | SCORE > 0 | 0.587 | 0.470 | 0.714 | 0.640 | 0.542 |
|  | **SCORE > -0.5** | **0.584** | **0.601** | **0.566** | **0.600** | **0.601** |
|  | SCORE > -1.0 | 0.578 | 0.728 | 0.417 | 0.574 | 0.642 |
|  | SCORE > -1.5 | 0.580 | 0.729 | 0.418 | 0.573 | 0.643 |

#### Impact of Clinical Note Integration on MASLD Phenotyping

Table 3 compares the performance of the selected CP rules with and without the incorporation of clinical notes for identifying MASLD in patients with T2D. According to the results, we can observe that integrating clinical notes consistently improved overall identification performance, with the largest gains observed in sensitivity and F1-score. Sensitivity increased by approximately 9-13%, while accuracy improved by 3-7% across the evaluated CP rules. Among the liver enzyme-based rules, ALT exhibited one of the largest performance improvements. Accuracy increased from 0.683 to 0.727, sensitivity from 0.705 to 0.791, and the F1-score from 0.696 to 0.749 following note integration. Similarly, the AST rule improved from 0.637 to 0.685 in accuracy, with sensitivity increasing from 0.665 to 0.762 and the F1-score from 0.653 to 0.713. The FNI rule achieved the highest sensitivity, increasing from 0.751 to 0.831, while maintaining comparable specificity (0.619 vs. 0.616). The corresponding F1-score also increased from 0.705 to 0.750. Similar improvements were observed for FIB-4 and the NAFLD Score, with gains in sensitivity, PPV, and F1-score accompanied by only minor reductions in specificity. Overall, incorporating clinical note-derived evidence consistently improved the detection of MASLD across all rules while largely preserving specificity, demonstrating the complementary value of unstructured clinical data for rule-based EHR phenotyping.

**Table 3.**
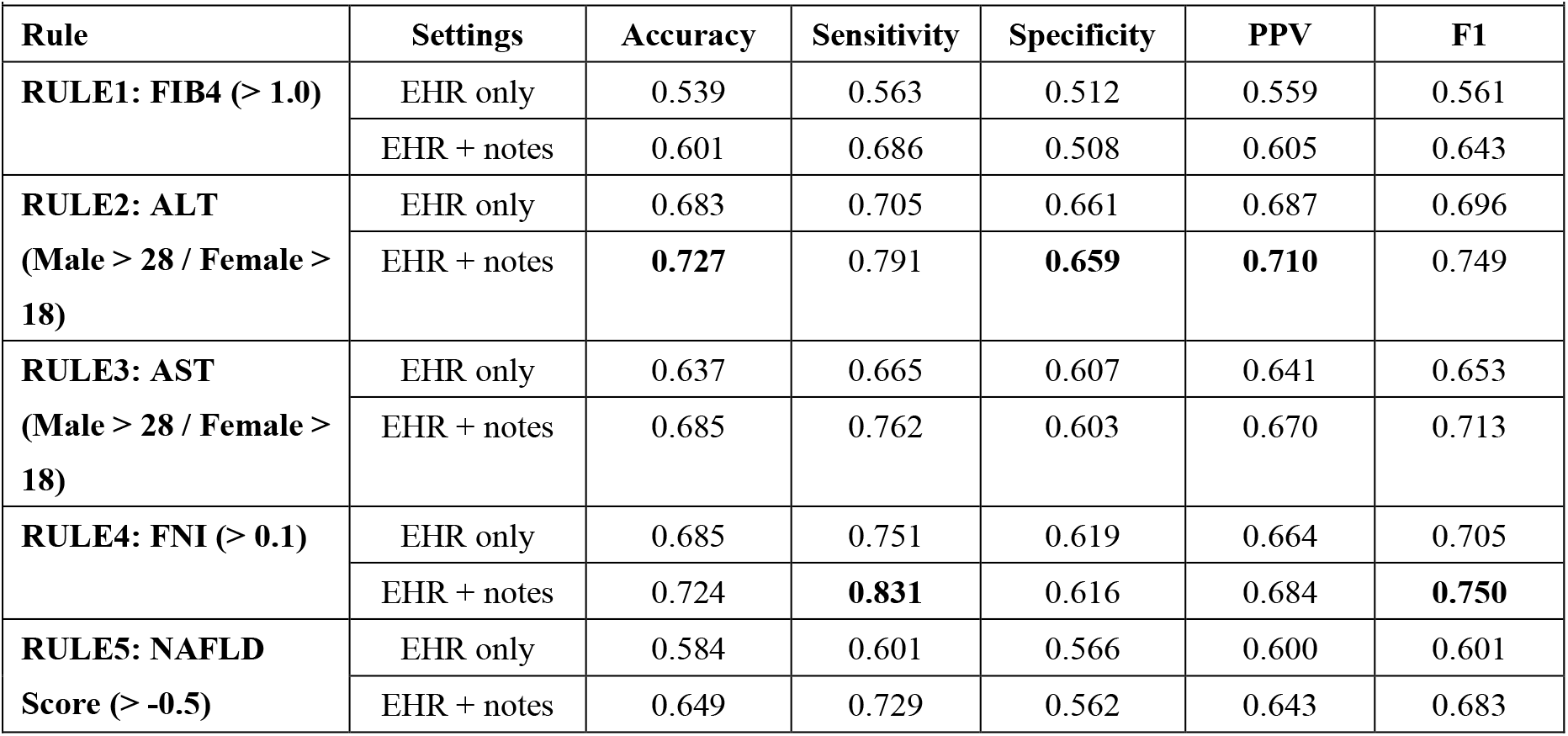
Comparison of the diagnostic performance of the selected rule-based CPs for MASLD using structured EHR data alone and after integration of clinical note-derived evidence.

| Rule | Settings | Accuracy | Sensitivity | Specificity | PPV | F1 |
| --- | --- | --- | --- | --- | --- | --- |
| RULE1: FIB4 (> 1.0) | EHR only | 0.539 | 0.563 | 0.512 | 0.559 | 0.561 |
|  | EHR + notes | 0.601 | 0.686 | 0.508 | 0.605 | 0.643 |
| RULE2: ALT<br>(Male > 28 / Female > 18) | EHR only | 0.683 | 0.705 | 0.661 | 0.687 | 0.696 |
|  | EHR + notes | **0.727** | 0.791 | **0.659** | **0.710** | 0.749 |
| RULE3: AST<br>(Male > 28 / Female > 18) | EHR only | 0.637 | 0.665 | 0.607 | 0.641 | 0.653 |
|  | EHR + notes | 0.685 | 0.762 | 0.603 | 0.670 | 0.713 |
| RULE4: FNI (> 0.1) | EHR only | 0.685 | 0.751 | 0.619 | 0.664 | 0.705 |
|  | EHR + notes | 0.724 | **0.831** | 0.616 | 0.684 | **0.750** |
| RULE5: NAFLD<br>Score (> -0.5) | EHR only | 0.584 | 0.601 | 0.566 | 0.600 | 0.601 |
|  | EHR + notes | 0.649 | 0.729 | 0.562 | 0.643 | 0.683 |

### Subgroup Performance Analysis

#### Age-Stratified Performance

Figure 2 summarizes the performance of the five CP rules across different age groups. The age-dependent rules, FIB-4 and the NAFLD Score, exhibited distinct age-related trends, with sensitivity increasing and specificity decreasing as age increased. Despite these opposing trends, overall accuracy remained relatively stable across age strata. In contrast, the liver enzyme-based (ALT and AST) and FNI rules demonstrated substantially more consistent performance across age groups. Their sensitivity, specificity, and accuracy varied only modestly, with overall accuracy generally ranging from approximately 0.68 to 0.75. Among these rules, FNI exhibited the most stable performance across all age groups, whereas ALT and AST showed only minor fluctuations. These findings suggest that age-dependent scoring systems are more sensitive to changes in patient age, whereas laboratory-based rules maintain relatively consistent performance across different age populations.

**Figure 2.**
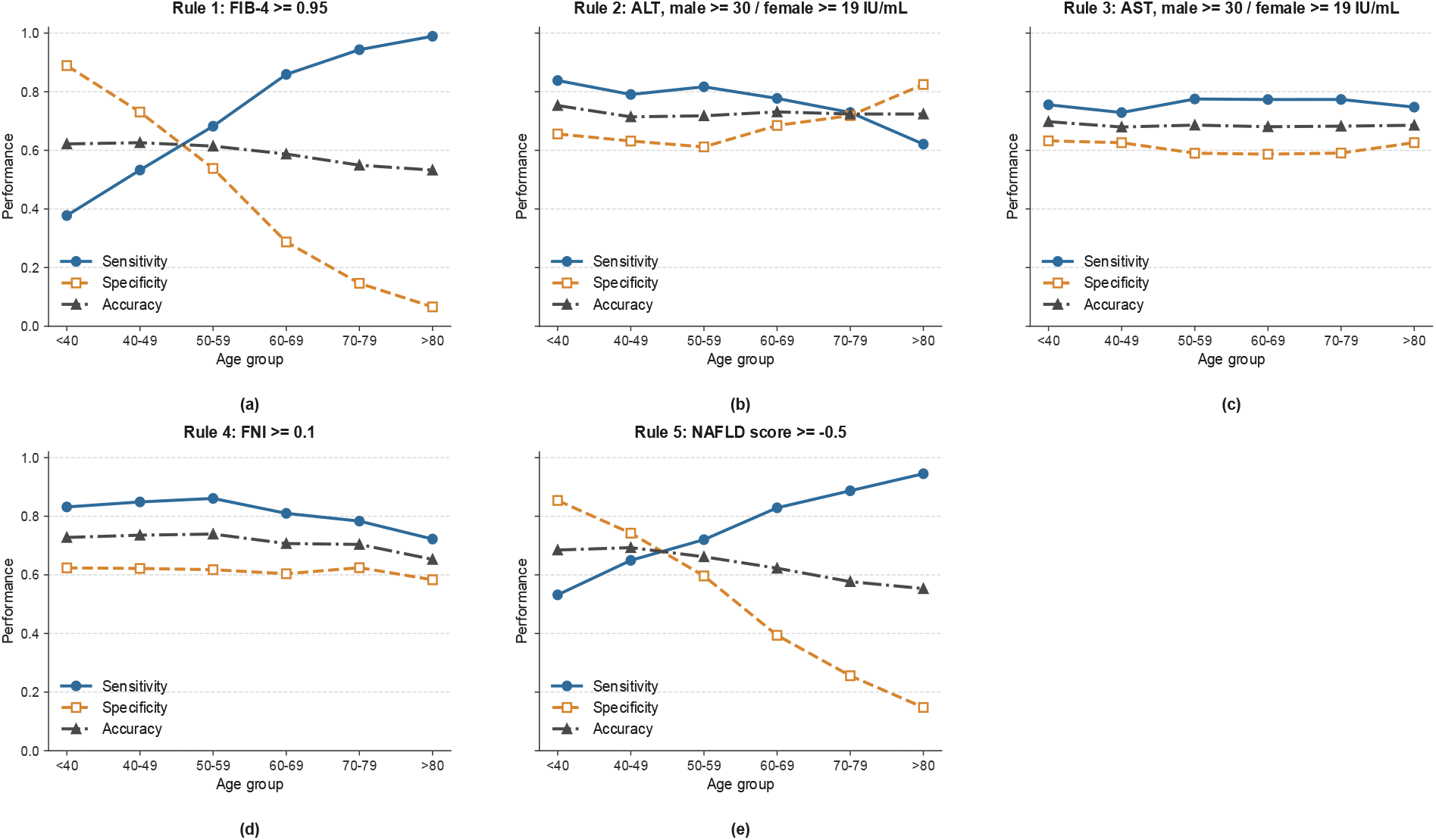
Age-stratified performance across five computable phenotype rules. Lines show sensitivity, specificity, and accuracy within each age group.

#### Sex-Stratified Performance

Figure 3 illustrates the sex-stratified performance of identifying MASLD using the proposed CP framework. Among the liver enzyme-based rules, ALT demonstrated similar performance in both sexes, with females showing slightly higher sensitivity accompanied by a modest reduction in specificity. A similar trend was observed for AST, although the difference between sensitivity and specificity was more pronounced in females. The FNI rule maintained high sensitivity in both subgroups, while females exhibited moderately higher specificity and overall accuracy than males. In contrast, FIB-4 and the NAFLD Score demonstrated only minor variations between sexes, with largely comparable diagnostic performance. Overall, the sex-stratified analysis indicates that the selected CP rules are robust across male and female populations, with only modest shifts in the sensitivity-specificity balance and no substantial differences in overall accuracy.

**Figure 3.**
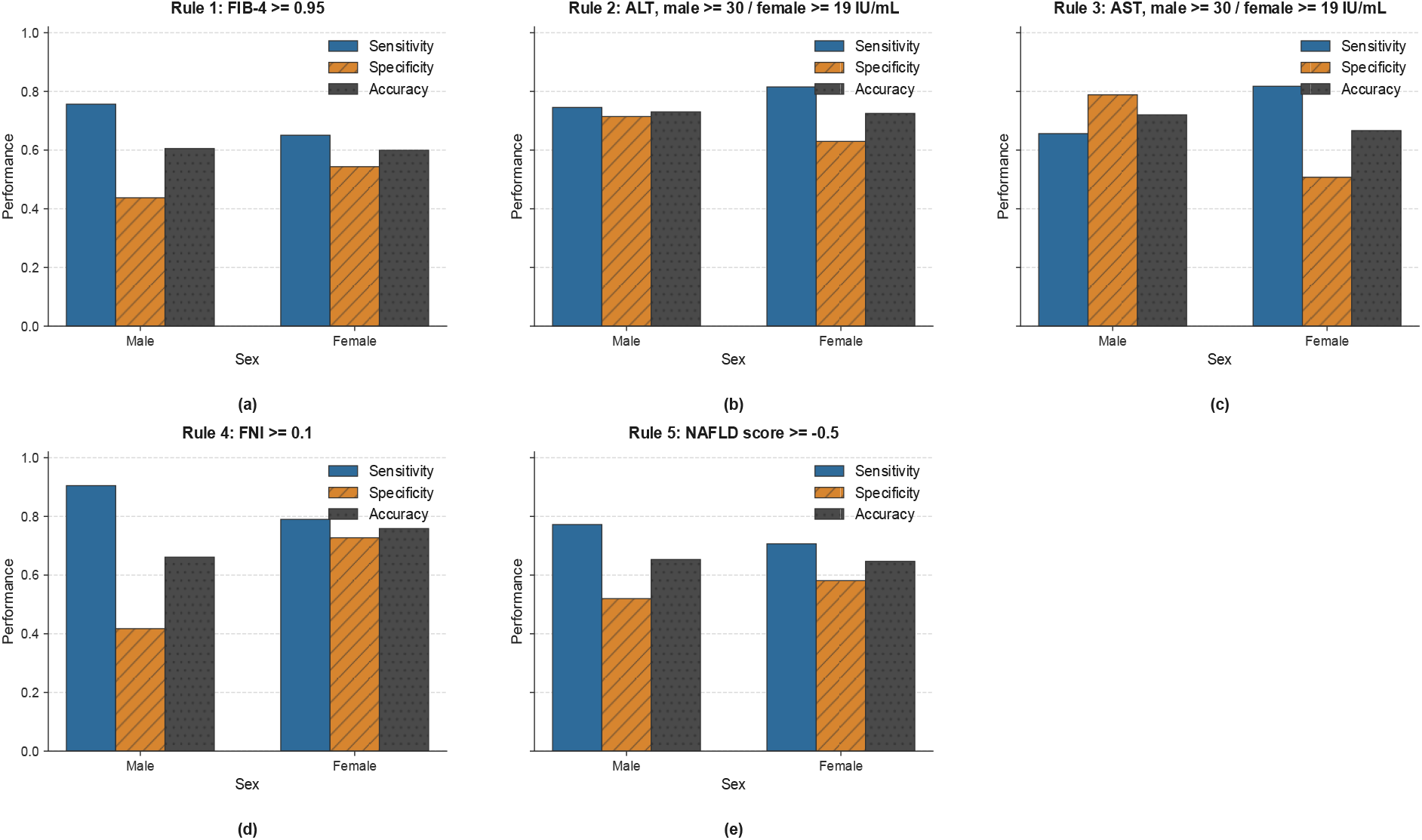
Sex-stratified performance across five rules. Bars report sensitivity, specificity, and accuracy for male and female subgroups.

#### Validation Against Manual Chart Review

To further assess the validity of the proposed CP framework, a subset of patient records identified from the UF Health T2D cohort was randomly selected for expert manual chart review. After eligibility screening and quality assessment, 240 patient records received definitive manual MASLD classifications. The expert review incorporated all available demographic information, laboratory measurements, and clinical documentation, providing an independent clinical assessment of MASLD status. Manual chart review confirmed an accuracy of 0.803, sensitivity of 0.709, specificity of 0.924, PPV of 0.922, and an F1-score of 0.802. The high specificity and PPV indicate that most patients classified as MASLD by the proposed framework were confirmed through expert review, whereas the comparatively lower sensitivity suggests that a subset of manually identified MASLD cases was not captured by the rule-based approach. Overall, the chart review findings provide independent clinical support for the validity of our proposed framework for identifying MASLD from EHR data.

## Discussion

This study developed and validated a rule-based and data-driven CP framework for identifying MASLD in patients with T2D, integrating both structured EHR and unstructured clinical notes. Overall, the proposed CP framework exhibited robust performance and consistency across multiple CP rules and demographic subgroups, supporting its utility for real-world MASLD phenotyping. Among the evaluated rules, the ALT-, AST-, and FNI-based CP rules achieved the most balanced performance, while the incorporation of clinical note-derived information consistently improved sensitivity and F1-score with minimal changes in specificity. These findings demonstrate that structured and unstructured EHR data provide complementary information for improving MASLD case identification in real-world clinical settings.

Several methodological advantages distinguish the proposed framework from existing EHR-based MASLD phenotyping approaches. First, multiple clinically derived phenotyping rules were systematically evaluated across a range of threshold values rather than relying exclusively on guideline-recommended cutoffs, enabling selection of parameters better suited to real-world EHR data. Second, unstructured clinical notes were incorporated through a lightweight, interpretable NLP pipeline that combined a curated MASLD terminology lexicon with contextual filtering, including negation detection and exclusion of family history. This approach captured clinically relevant evidence frequently absent from structured fields and consistently improved case detection. Finally, we incorporated propensity score matching to reduce demographic imbalance and performed it at the encounter level, enabling assessment across longitudinal clinical observations. Together, these components provide a scalable and reproducible EHR-based CP framework for identifying MASLD in patients with T2D.

Beyond MASLD identification, this study highlights the value of integrating complementary EHR modalities within CP development. Many chronic diseases are incompletely represented by structured diagnosis codes or laboratory measurements alone, whereas clinically important diagnostic evidence is often documented in free-text narratives. The observed improvement in sensitivity following note integration demonstrates that routine clinical documentation can meaningfully enhance computable phenotype performance while preserving interpretability. The proposed framework therefore has potential applications in large-scale cohort identification, real-world evidence generation, epidemiologic studies, and clinical trial recruitment, and its modular design can be adapted to phenotyping other chronic diseases that similarly rely on heterogeneous EHR data sources.

This study has several limitations. First, the note-processing component relied on keyword matching with contextual filtering and therefore may not fully capture the semantic complexity of clinical narratives. Although negation detection and context filtering reduced false-positive matches, more advanced NLP approaches, such as contextual language models or transformer-based clinical NLP systems, may further improve contextual understanding and reduce misclassification. Second, validation was limited to manual review of 240 records from a single institution. External validation using multicenter EHR datasets will be necessary to evaluate the portability and generalizability of the proposed framework. Third, the selected rule thresholds were optimized using the current study population and may require recalibration in healthcare systems with different patient characteristics, documentation practices, or laboratory distributions. Future work will investigate adaptive threshold selection, incorporate longitudinal laboratory trajectories, and explore more advanced NLP methods to further improve CP performance.

## Conclusion

This study presents a rule-based CP framework for identifying MASLD in patients with T2D by integrating structured EHR data with information extracted from unstructured clinical notes. The proposed framework demonstrated robust and consistent diagnostic performance across multiple rule-based algorithms, with clinical note integration further improving case identification while maintaining stable performance across demographic subgroups. Validation through expert manual chart review provided additional clinical support for the reliability of the proposed approach. By combining interpretable rule-based algorithms with lightweight natural language processing, the framework preserves both transparency and scalability, making it well suited for large-scale real-world EHR studies. Beyond MASLD, the proposed methodology provides a practical and generalizable strategy for developing multimodal EHR-based CPs for other chronic diseases and supports future applications in clinical research, population health surveillance, and real-world evidence generation.

## Data Availability

The data analyzed in this study were obtained from the University of Florida Health Integrated Data Repository (IDR). Due to institutional policies and patient privacy considerations, the underlying patient-level data are not publicly available. Access to the data may be considered upon reasonable request and subject to applicable institutional approvals and data access requirements.

## Acknowledgement

We acknowledge the University of Florida Integrated Data Repository (IDR) and the UF Health Office of the Chief Data Officer for providing the analytic data set for this project. Additionally, the research reported in this publication was supported by the National Center for Advancing Translational Sciences of the National Institutes of Health under the University of Florida Clinical and Translational Science Award UL1TR001427.

